# Cost-effectiveness of Ezetimibe plus statin lipid-lowering therapy: A systematic review and meta-analysis of cost-utility studies

**DOI:** 10.1101/2022.02.14.22270957

**Authors:** Akhil Sasidharan, S Sajith Kumar, Kayala Venkata Jagadeesh, Meenakumari Natarajan, Bhavani Shankara Bagepally

## Abstract

**Introduction:** In addition to statin therapy, Ezetimibe, a non-statin lipid-modifying agent, is increasingly used to reduce low-density lipoprotein cholesterol and atherosclerotic cardiovascular disease risk. Literature suggests the clinical effectiveness of Ezetimibe plus statin (EPS) therapy; however, primary evidence on its economic effectiveness is inconsistent. Hence we pooled incremental net benefit to synthesise the cost-effectiveness of EPS therapy.

**Methods:** We identified economic evaluation studies reporting outcomes of EPS therapy compared with other lipid-lowering therapeutic agents or placebo by searching PubMed, Embase, Scopus, and Tufts Cost-Effective Analysis registry. Using random-effects meta-analysis, we pooled Incremental Net Benefit (INB) in the US $ with a 95% confidence interval (CI). We used the modified economic evaluations bias checklist and GRADE quality assessment for quality appraisal. The review was apriori registered with PROSPERO, CRD42021248531.

**Results:** The pooled INB from twenty-one eligible studies showed that EPS therapy was significantly cost-effective compared to other lipid-lowering therapeutic agents or placebo. The pooled INB (95% CI) was $4,274 (621 to 7,927), but there was considerable heterogeneity (I^2^=84.21). On subgroup analysis EPS therapy is significantly cost-effective in high-income countries [$4,356 (621 to 8,092)], for primary prevention [$4,814 (2,523 to 7,106)], and for payers’ perspective [$3,255 (571 to 5,939)], and from lifetime horizon [$4,571 (746 to 8,395)].

**Conclusion:** EPS therapy is cost-effective compared to other lipid-lowering therapeutic agents or placebo in high-income countries and for primary prevention. However, there is a dearth of evidence from lower-middle-income countries and the societal perspective.

## Introduction

Cardiovascular disease (CVD) and cardiovascular events have a perpetual relationship with hyperlipidemia ^1-3^. The World Health Organization (WHO) reported that an estimated 17.9 million people died from CVDs in 2019 alone, representing one-third of all global deaths ^4^. For reducing cardiovascular events, statin drugs targeting 3-hydroxy-3-methyl-glutaryl-coenzyme (HMG-CoA) reductase are the most prescribed medications ^5^. Statins lower the cardiovascular risk in all groups ^6-8^, as well as the risk of developing CVD events ^9, 10^. Despite rigorous statin regimens aimed to lower the risk of cardiovascular complications, a large number of statin-treated patients fail to attain the recommended target low-density lipoprotein-cholesterol (LDL-C) levels due to statin intolerance or discontinuation of treatment due to adverse drug reactions ^11, 12^. Ezetimibe is a non-statin lipid-modifying agent targeting the Niemann-Pick C1-like 1 intestinal cholesterol transporter protein (cholesterol absorption inhibitor) ^13, 14^. It is used to achieve the desirable LDL-C levels for patients on maximally tolerated statin therapy. When added to a statin, Ezetimibe achieves a reduction in LDL-C of typically 20–25% with reduced atherosclerotic CVD (ASCVD) risk ^15, 16^. Hence, the 2018 Cholesterol guidelines warrant using Ezetimibe in individulas with a high ASCVD risk despite receiving optimal statin medication ^7^. Studies showed that Ezetimibe reduced LDL-C at levels consistent with Cholesterol Treatment Trialists’ (CTT) Collaboration estimates, giving CTTs’ extrapolation beyond their intial analysis legitimacy and validity ^15, 17^. Many professional organisations have recently issued guidelines recommending the use of non-statin medications in clinical practice, consideringtheir usefulness ^3, 18-20^.

Ezetimibe co-administration with statins has resulted in fewer very-high-risk and extremely high-risk patients eligible for other lipid-lowering agents ^21^. A favourable tolerability profile, ease of use, and affordability make Ezetimibe a better option than PCSK9i ^22^. Furthermore, a recent meta-analysis of cost-utility studies (CUA) showed PCSK9i to be not cost-effective compared to other lipid-lowering therapeutic agents in high-income countries (HICs) ^23, 24^. As a result, Ezetimibe can be used as a next-cholesterol medication. However, its use and the extent to which it meets unmet reuirements are limited. The availability of Ezetimibe as a generic drug in several countries ^25^ can act as a positive indicator and increase overall access to this medication.

Further, the evidence on the cost-effectiveness of this therapy is also inconsistent, as few studies report it is cost-effective ^26 27^. In contrast, other studies report that combination therapy is not cost-effective^20 28^ compared to statin therapy. Hence to provide syntheised evidence, we systematically reviewed the evidence on the cost-effectiveness and quantitatively estimated the pooled incremental net benefit (INB) of ezetimibe therapy.

## METHODS

The study protocol was pre-registered with PROSPERO, CRD42021248531, and the study was conducted according to the Preferred Reporting Items for Systematic Reviews and Meta-analyses (PRISMA) ^29^.

### Data sources, eligibility criteria, screening, and search strategy

We searched PubMed, Embase, Scopus, and the Tufts Medical Centers’ cost-effective analysis (CEA) registry^30^ for studies published from inception to 26 April 2021 (see Appendix 1), adhering to the PRISMA guideline. We followed the PICO approach (Population, Intervention, Comparator, Outcome) to construct the search terms. Published Cost-Utility Analysis (CUAs) were eligible if they met all the following inclusion criteria. Adult subjects (age above 18 years) with risk of or established CVD requiring lipid-lowering therapy and treated with Ezetimibe compared to other lipid-lowering therapeutic agents such as statins or PCSK9i, or with placebo/no treatment. We included studies reporting economic outcomes in incremental cost-effectiveness ratios (ICER) per quality-adjusted life years (QALYs) or INB. Studies with effectiveness measured other than in quality-adjusted life years (QALY), reviews, letters, editorials, abstracts, books, reports, grey literature, and methodological articles were excluded. The detailed search terms are reported in Appendix 1.

### Selection of studies

We identified a total of 1,944 studies after systematically searching multiple peer-reviewed repositories. All English language studies that met the eligibility criteria listed from the electronic database search were screened independently for titles and abstracts by two independent reviewers (BSB and AS) for their potential inclusion using the Rayyan-web application ^31^. Reviewers (AS, KVJ, and MK) independently reviewed the full text of the finalized 125 studies after the title and abstract screening and deduplication in detail. The independent assessors’ mutual agreement with another reviewer (BSB) produced the final list of studies meeting the inclusion and exclusion criteria (n=22), and data were extracted from the selected studies. The PRISMA flow chart of the screening process is appended as Figure 1.

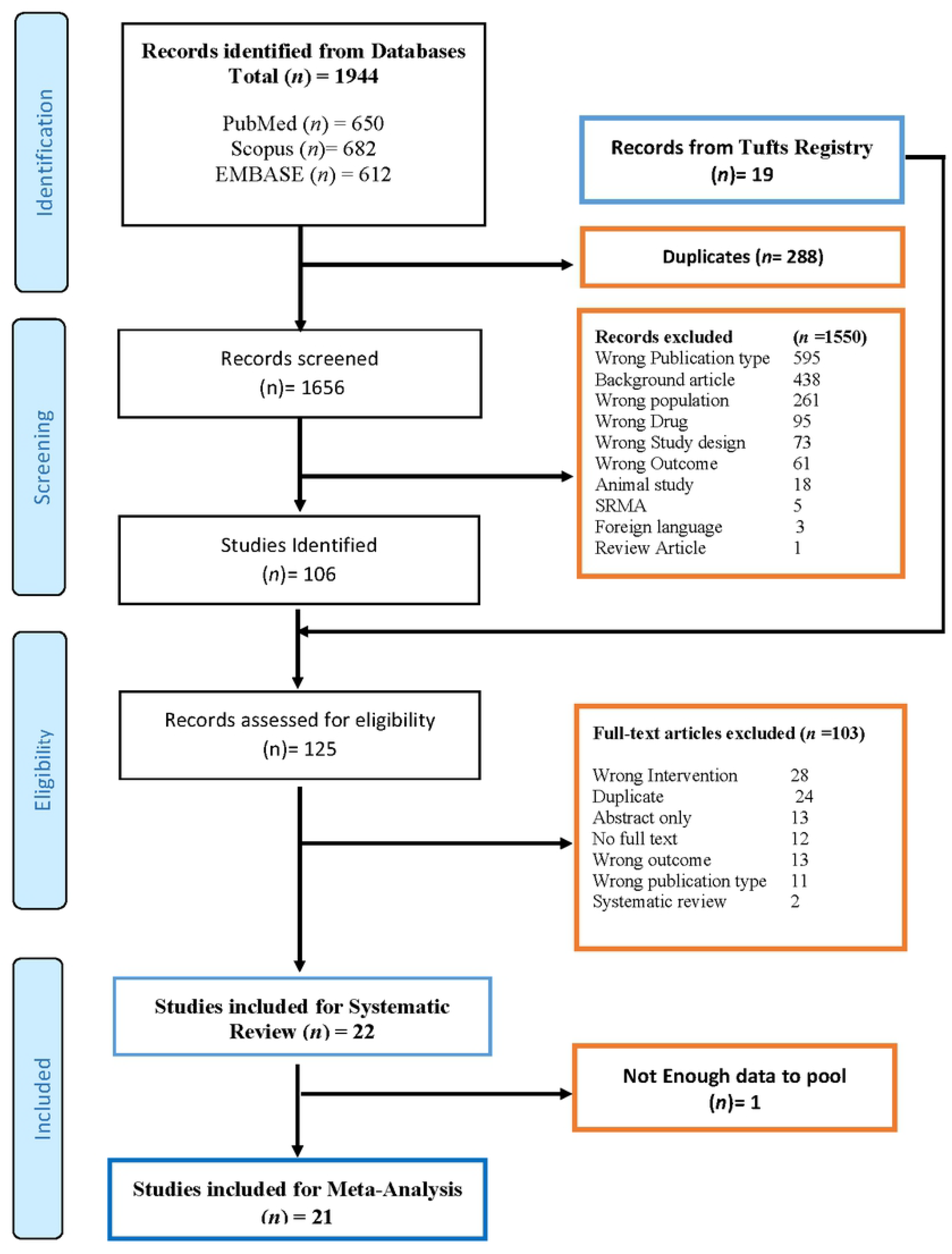

### Data extraction

Using a data extraction template adapted for the outcomes of interest, we extracted the following data from eligible studies: author, year, country of setting, study/patient characteristics, intervention, comparator, and the general characterization of the model, which included model type, perspective, time horizons, discount rate, and currency year. We extracted economic parameters such as costs (C), incremental costs (ΔC), clinical effectiveness (E), its incremental effectiveness (ΔE), ICERs, INB values, and their measures of dispersion [i.e., standard deviation (SD), standard error (SE), or 95% confidence interval (CI), willingness to pay (WTP), and threshold (K). From the cost-effective (CE) plane graph, we extracted ΔC and ΔE values using WebPlotDigitaliser ^32^.

### The outcome of interest

In the meta-analysis, we estimated the pooled INB, defined as pooled INB = K*ΔE-ΔC, where K was the WTP threshold, ΔC-incremental cost (i.e., the difference in costs between intervention and comparator), ΔE-incremental effectiveness (i.e., the difference in effectiveness between intervention and comparator). A positive INB favours intervention, i.e., intervention is cost-effective, whereas a negative INB favours the comparator, i.e., intervention is not cost-effective. INB is used instead of ICER as the effect measure because of the inherent limitations of ICER and the ambiguity in interpreting them ^33-35^.

### Data preparation and statistical analysis

We followed the data preparation method and analysis described and used elsewhere ^23, 33, 36, 37^. To calculate the INB and its variance, mean values along with dispersions (SD, SE, and 95% CI) of ΔC and ΔE are required. However, economic studies reported different parameters; therefore, we designed five scenarios to deal with the data available from primary studies (Appendix II). Using the data reported in the primary research publications and following the approach detailed in Bagepally et al. ^33, 36^, we calculated the INB and its variances for each intervention comparator duo. If a cost-effective (CE) plane graph was not provided, covariance was estimated for 1000 Monte-Carlo simulations from the extracted ΔC and ΔE values for studies included in scenario 3 (see Appendix II).

Included studies reported in different currencies from different time points (years). To compare INB in a common currency, all monetary units, except for the non–GDP-based threshold, were adjusted for inflation using the consumer price index (CPI) and converted to purchasing power parities (PPP)-adjusted US dollar (US $) for the year 2021, as detailed in Appendix II. Following the data preparation, INBs were pooled across studies stratified by income classification as low-income (LIC), lower-middle (LMIC), upper-middle (UMIC), and high-income (HIC) countries as per the World Bank classification ^38^. Meta-analysis was applied to pool the INBs using random-effects model based on the DerSimonian and Laird method. I^2^ statistics were used to assess heterogeneity, I^2^ > 50% was considered substantial heterogeneity, and Cochrane Q p-value < 0.05 was taken as a cut-off for significant heterogeneity. We did subgroup analysis wherever appropriate to explore the source of heterogeneity and provide subgroup-specific pooled INBs. Subsequently, we assessed the publication bias using funnel plots and Eggers’ test. Furthermore, we explored the sources of asymmetry using contour-enhanced funnel plots. All data were prepared using Microsoft Excel version 2019 ^39^ and analyzed using Stata software version 16 ^40^.

### Risk of bias assessment and quality assessment

Reporting quality was assessed independently by the reviewers using the modified economic evaluation bias (ECOBIAS) checklist ^41^. It considers both overall biases (11 items) and model-specific biases, including structure (4 items), data (6 items), and internal consistency (1 item). Each item was rated as applicable, partially applicable, unclear, no, or not applicable (Supplementary Figure 1). GRADE (Grading of Recommendation, Assessment, Development, and Evaluation) was used to assess the quality of evidence and grade recommendations ^42, 43^. We graded the evidence for the cost-effectiveness of Ezetimibe plus statin (EPS) therapy compared to other lipid-lowering therapeutic agents or a placebo. The certainty of the evidence was rated for EPS therapy in HICs, for primary prevention, from a payer’s perspective, and over a lifetime horizon. Additionally, the certainty of the evidence was rated for the cost-effectiveness of EPS therapy versus statin monotherapy, for EPS therapy versus statin monotherapy for primary prevention from the payer’s perspective as well. This assessment was based on the risk of bias, inconsistency, indirectness, imprecision, publication bias, and other considerations. The quality of the evidence was classified as high, moderate, low, or very low ^42, 43^. We excluded study design and sample size considerations because the included cost-utility studies are model-based.

## Results

### General Characteristics of the Included Studies

We retrieved and included twenty-two potentially relevant articles^20, 26-28, 44-61^ for systematic review, of which 21 studies were eligible for meta-analysis^20, 26-28, 44-46, 48-61^ (Figure 1). One study from the UK, Ara et al. ^47^, was not included for meta-analysis due to incomplete data. The general characteristics of the included articles are summarised in Table 1. Included studies reported 25 comparisons; EPS therapy versus statin monotherapy(n=18)^26-28, 44, 46, 48-50, 52-55, 58-61^, EPS therapy versus PCSK9i plus Ezetimibe and statins(n=3)^20, 56, 57^, EPS therapy versus PCSK9i with statin therapy(n=2)^44, 60^ and EPS therapy versus no treatment or placebo(n=2)^45, 51^. All studies were set in HICs, except two, based on UMICs^28, 59^. Most of the studies (n = 20) analysed a lifetime horizon, except two studies^51, 59^. Fourteen studies reported from a healthcare perspective^26, 28, 44-46, 51, 53-56, 58-60^, four from payer’s perspective^20, 27, 48, 52^, and three studies from a societal perspective^28, 49, 50^. All studies used Markov models with acycle length of one year, except for an alongside trial^51^. The named models included were Cardio Vascular Disease Policy Model (CVDPM) (n = 3)^44, 53, 60^ and the COOK model (n = 2)^26, 52^. All studies used discounting for cost and outcomes. All of the included studies reported direct medical costs; besides, Kongpakwattana et al. reported direct non-medical costs^28^, and Landmesser et al. reported indirect costs^20^. All the studies reported using country-specific thresholds as WTP except one study^59^ that used GDP-based WTP.

**Table 1.** – Characteristics of identified studies for Systematic Review

| Author_year | Country | Setting | Perspective | Target population | Time Horizon | Discount Rate | Reference year | Intervention | Comparator | Prevention | Remarks |
| --- | --- | --- | --- | --- | --- | --- | --- | --- | --- | --- | --- |
| Kohli_2006<sup>26</sup> | Canada | Risk Group | Healthcare | CAD & non-fatal CAD (angina or acute MI) | Lifetime | 5 | 2002 | Ezetimibe+ Statin | Statin | PP+SP | Cost effective |
| Ara_2008<sup>1 45</sup> | UK | Country | Healthcare | Primary Hypercholesterolemia | Lifetime | 3.5 | 2006 | Ezetimibe+ Statin | No Treatment | PP | Cost effective |
| Ara_2008<sup>2 46</sup> | UK | Risk Group | Healthcare | Primary Hypercholesterolemia | Lifetime | 3.5 | 2006 | Ezetimibe+ Statin | Statin | PP | Cost effective |
| Ara_2008<sup>3 47*</sup> | UK | Risk Group |  | ACS | Lifetime | 3.5 | 2006 | Ezetimibe+ Statin | Statin |  | Cost effective |
| Reckless_2010<sup>48</sup> | USA | Risk group | Payer | Non-fatal CHD with or without DM | Lifetime | 3.5 | 2004 | Ezetimibe+ Statin | Statin | SP | Cost effective |
| Soini_2010<sup>49</sup> | Finland | Risk group | Societal | Primary Hypercholesterolemia | Lifetime | 3 | 2007 | Ezetimibe+ Statin | Statin | SP | Cost effective |
| Nooten_2011<sup>50</sup> | Netherland | Country | Societal | High CVD risk, history of CHD and/or DM | Lifetime | 4 | 2008 | Ezetimibe+ Statin | Statin | PP | Cost effective |
| Laires_2015<sup>27</sup> | Portugal | Country | Payer | CKD but without known CHD | Lifetime | 5 | 2015 | Ezetimibe+ Statin | Statin | PP | Cost effective |
| Mihaylova_2016<sup>51</sup> | UK | Country | Healthcare | HeterozygousFH/ Preexisting ASCVD | Non-Lifetime | 3.5 | 2015 | Ezetimibe+ Statin | No Treatment | PP | Not Cost effective |
| Kazi_2016<sup>a 44</sup> | USA | Country | Healthcare | HeterozygousFH/ Preexisting ASCVD | Lifetime | 3 | 2015 | Ezetimibe+ Statin | Statin | SP | Cost effective |
| Kazi_2016<sup>b 44</sup> | USA | Country | Healthcare | ASCVD | Lifetime | 3 | 2015 | Ezetimibe+ Statin | PCSK9i+ Statin | SP | Cost effective |
| Kazi_2017<sup>53</sup> | USA | Risk Group | Healthcare | ACS | Lifetime | 3 | 2017 | Ezetimibe+ Statin | Statin | SP | Cost effective |
| Almalki_2017<sup>55</sup> | Saudi Arabia | Country | Healthcare | CVD history (both CHD and stroke). | Lifetime | 3 | 2016 | Ezetimibe+ Statin | Statin | SP | Cost effective |
| Davies_2017<sup>52</sup> | USA | Country | Payer | CAD & non-fatal CAD (angina or acute MI) | Lifetime | 3 | 2013 | Ezetimibe+ Statin | Statin | PP+SP | Cost effective |
| Stam-Slob_2017<sup>54</sup> | Netherland | Risk group | Healthcare | stable CAD | Lifetime | 4 | 2014 | Ezetimibe+ Statin | Statin | SP | NA |
| Korman_2018<sup>61</sup> | Norway | Country | Healthcare | Hypercholesterolemia, DM, Statin Intolerant | Lifetime | 4 | 2015 | Ezetimibe+ Statin | Statin | PP+SP | Cost effective |
| Stam-Slob_2018<sup>56</sup> | Netherland | Risk group | Healthcare | FH without a history of vascular disease, stable vascular disease, DN | Lifetime | 3 | 2014 | Ezetimibe+ Statin | PCSK9i+ Ezetimibe+ Statin | SP | NA |
| Kongpakwattana_2019<sup>a 28</sup> | Thailand | Country | Societal | Existing CVD, comprising MI and stroke | Lifetime | 3 | 2018 | Ezetimibe+ Statin | Statin | SP | Not Cost effective |
| Kongpakwattana_2019<sup>b 28</sup> | Thailand | Country | Healthcare | Existing CVD, comprising MI and stroke | Lifetime | 3 | 2018 | Ezetimibe+ Statin | Statin | SP | Not Cost effective |
| Kazi_2019<sup>a 60</sup> | USA | Country | Healthcare | ACS | Lifetime | 3 | 2018 | Ezetimibe+<br>Statin | Statin | SP | Cost effective |
| Kazi_2019<sup>b 60</sup> | USA | Country | Healthcare | ACS | Lifetime | 3 | 2018 | Ezetimibe+<br>Statin | PCSK9i+<br>Statin | SP | Cost effective |
| Schlackow_2019<sup>a 58</sup> | USA | Risk group | Healthcare | Non dialysis patients with CKD | Lifetime | 3 | 2015 | Ezetimibe+<br>Statin | Statin | PP | Cost effective |
| Schlackow_2019<sup>b 58</sup> | UK | Risk group | Healthcare | Non dialysis patients with CKD | Lifetime | 3.5 | 2015 | Ezetimibe+<br>Statin | Statin | PP | Cost effective |
| Dressel_2019<sup>57</sup> | Germany | Country |  | Stable CAD | Lifetime | 3 | 2018 | Ezetimibe+<br>Statin | PCSK9i+<br>Ezetimibe+<br>Statin | SP | Not Cost effective |
| Landmesser_2020<sup>20</sup> | Sweden | Risk group | Payer | Recent MI /MI with a second event/MI with a risk factor | Lifetime | 3 | 2019 | Ezetimibe+<br>Statin | PCSK9i+<br>Ezetimibe+<br>Statin | SP | Not Cost effective |
| Han Yang_2020<sup>59</sup> | China | country | Healthcare | Newly diagnosed with CVD | Non-Lifetime | 3 | 2017 | Ezetimibe+<br>Statin | Statin | SP | Cost effective |

\*Not included in meta-analysis, PP-primary prevention, SP-secondary prevention, PP+SP- both primary and secondary prevention

In the meta-analysis, due to the differences in reported outcomes among different studies, the INB variance from the most comparable studies was utilised for calculations under scenario five (Appendix II). The INB variance of Van Nooten et al. ^50^ was used for three studies ^20, 49, 54^, and the INB variance of Schlackow et al. ^58^ was used for six studies ^26, 27, 45, 46, 51, 55^, respectively. Among the 21 studies evaluating the cost-effectiveness of Ezetimibe versus other lipid-lowering therapeutic agents or placebo, seven were set in Europe ^20, 27, 49, 50, 56, 57, 61^; five were set in the US ^44, 48, 52, 53, 58, 60^, three in the UK ^46, 47, 51^ and one studies each from Canada ^26^, China ^59^, Saudi Arabia ^55^ and Thailand ^28^. Schlackow et al. reported from the health system perspective of both the US and UK ^58^. In all studies, except Kongpakwattana et al. ^28^ and Almalki et al. ^55^ mean plasma LDL-C levels less than70 mg/dL reflected current ezetimibe dosing recommendations. Except for two^48, 49^ that included fatal and non-fatal stroke as well, all studies included fatal and non-fatal coronary heart diseases in their model. Unstable angina was modelled in three studies^48, 49, 55^ in which Almalki et al.^55^ considered coronary revascularisation also. Eventhough three studies^44, 53, 60^ modelled adverse events as consequences, only Yang et al.^59^ included costs due to adverse events. Six studies profiled the model population after local registries and databases^44, 49, 52, 53, 60, 61^, three studies ^48, 55, 59^ from clinical trials, only Kongpakwattana et al. used data from a meta-analysis of RCTs^28^ (see Table 1).

All three studies from payers’ perspective ^27, 48, 52^, and seven studies out of nine from the healthcare perspective ^26, 28, 46, 53-55, 58-60^ and two out of three studies from the societal perspective ^49, 50^ reported EPS therapy to be cost-effective compared with statin monotherapy. From the healthcare perspective, all two studies ^44, 60^ considered EPS therapy to be cost-effective compared to PCSK9i with statin therapy, but only one study^56^ out of three, reported EPS therapy to be cost-effective compared with PCSK9i along with Ezetimibe and statin. The alongside trial ^51^, which compared EPS therapy and placebo, reported that the intervention was not cost-effective. In contrast, a model-based study ^45^ that compared EPS therapy versus no treatment reported the intervention as cost-effective. In addition, according to the WTP threshold of Thailand, Kongpakwattana et al. ^28^ reported Ezetimibe as not cost-effective from healthcare or societal perspective. Three studies did not report any sensitivity analyses ^48, 53, 61^ while the remaining studies ^20, 26-28, 44-46, 49, 50, 52, 54-56, 59, 60^ includeddeterministic as well as probabilistic sensitivity analyses. Additionally, eight studies ^28, 44-46, 50, 54, 55, 60^ reported scenario and threshold analyses, and six studies ^26, 49, 51, 52, 56, 60^ also included sub-group analyses.

## 1. Quality Appraisal

### Risk of bias assessment

The risk of bias among the identified studies was analysed using the ECOBIAS checklist^62^. The ECOBIAS checklist shows that almost 91 percent of the studies chose the best current practice as a comparator, and all the comparators are adequately described. The details of the data used in the studies are transparent. All studies provided sufficient detail on the costs, effectiveness, discount rates, and have acknowledged te sources of of funding. Additionally, model selection bias was negligible. Similarly. bias related to time horizon was also low since majority of the studies employed a lifetime horizon. Limited scope bias is highly probable in almost all studies, also internal consistency related to mathematical logic was not evident (Supplementary Figure 1).

### Publication bias

The funnel plot showed asymmetry. The studies were distributed along with the mean effect size of the funnel plot. The Egger’s test with a higher p-value (p = 0.860) indicates no significant variability among the studies and no publication bias. However, the absence of studies in the area of significance on the contour enhanced funnel plot suggests the possibility of publication bias due to factors other than non-reporting bias (Supplementary Figure 2). Due to high heterogeneity, it would be difficult to distinguish between publication bias and other causes.

## 2. Ezetimibe compared with other lipid-lowering therapeutic agents or placebo

The pooled INB (INBp) with 95% CI, $4,274 (621 to 7,927) showed EPS therapy is significantly cost-effective compared with other lipid-lowering therapeutic agents or placebo. The INBp calculated from 25 comparisons^20, 26-28, 44-46, 48-61^ revealed considerable heterogeneity (I^2^ = 84.21) (Figure 2).

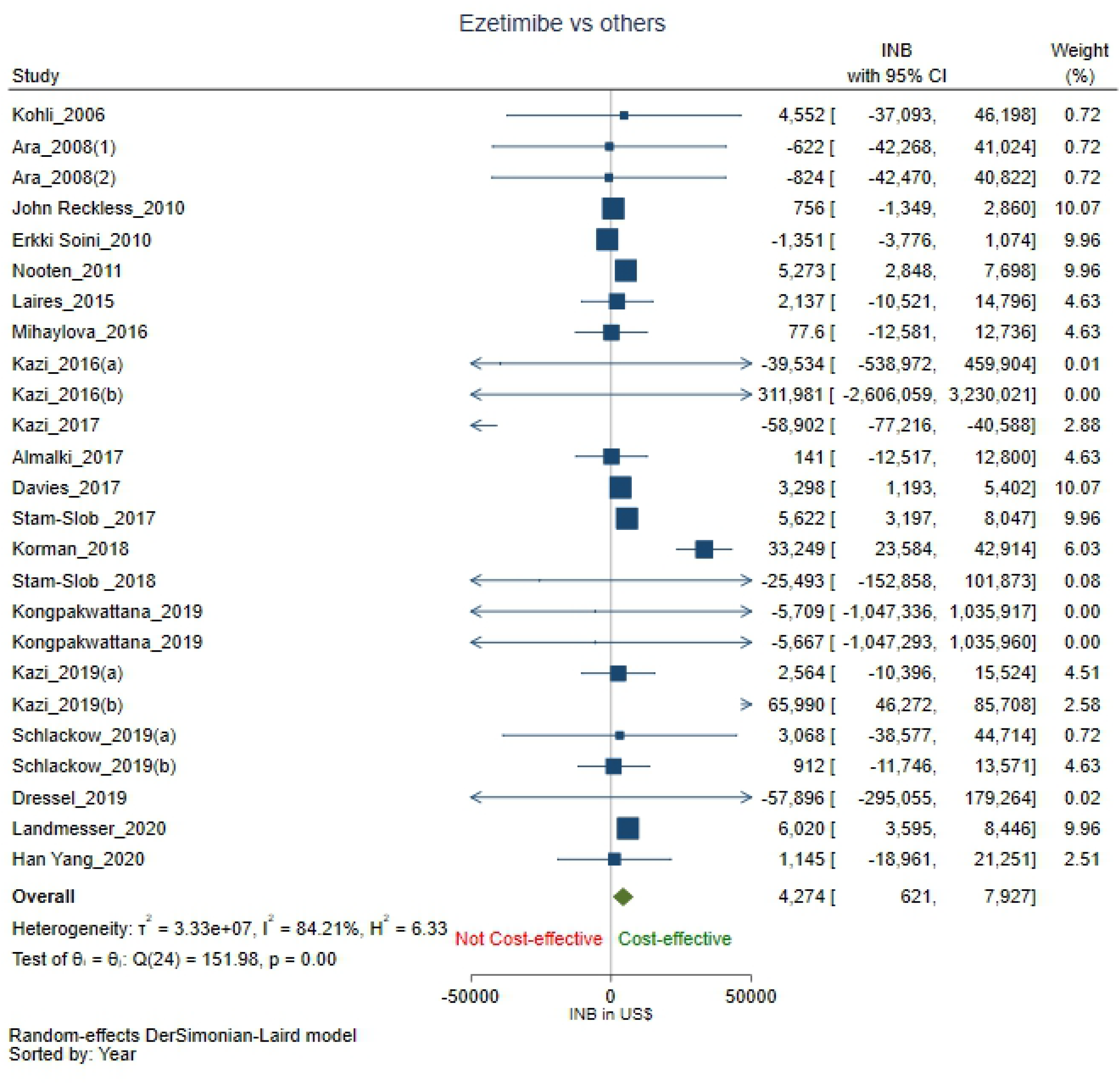

### Subgroup analysis

**We conducted** subgroup analyses to explore the heterogeneity between studies and provide subgroup specific pooled INBs. Subgroup analysis based on treatment comparisons showed that EPS therapy is significantly cost-effective compared with PCSK9i plus statin therapy (n = 2)^44, 60^ with INBp $66,001 (46,284 to 85,718). Also, EPS therapy is significantly cost-effective compared to PCSK9i plus EPS therapy (n = 3)^20, 56, 57^ with INBp $6,002 (3,578 to 8,427). There was no heterogeneity (I^2^ = 0.0) in either of the subgroups. However, EPS therapy is not significantly cost-effective compared to statin monotherapy (n = 18)^26-28, 44, 46, 48, 49, 52-55, 58-61^ with INBp $2,558 (−1,249 to 6,364) and considerable heterogeneity (I^2^ = 84.02). Likewise, EPS therapy is not significantly cost-effective compared to no treatment or placebo (n = 2)^45, 51^ with INBp $18.4 (−12,093 to 12,130) and no heterogeneity (I^2^ = 0.0) (Supplementary Figure 3).

On subgroup analysis with the income status of the countries, the pooled INBs from HICs (n = 22)^20, 26, 27, 44-46, 48-58, 60, 61^ showed that EPS therapy is cost-effective compared with other lipid-lowering therapeutic agents or placebo with an INBp of $4,356 (621 to 8,092) with considerable heterogeneity (I^2^ = 86.18). In contrast, EPS therapy is not significantly cost-effective for the UMICs^28, 59^ with an INBp of $1,140 (−18,959 to 21,239) and no heterogeneity (I^2^ = 0.0) (Supplementary Figure 4).

On subgroup analysis with study perspective, the pooled INBs among studies from payers’ perspective (n = 4)^20, 27, 48, 52^ showed that EPS therapy is significantly cost-effective compared with other lipid-lowering therapeutic agents or placebo with an INBp of $3,255 (571 to 5,939) but with substantial heterogeneity (I^2^ = 71.14). However, EPS therapy is not significantly cost-effective from a healthcare perspective (n = 17)^26, 28, 44-46, 51, 53-56, 58-61^ with INBp $3,255 (571 to 5,939) and considerable heterogeneity (I^2^ = 86.14) or from a societal perspective (n = 3)^28, 49, 50^ with INBp $3,255 (571 to 5,939) and considerable heterogeneity (I^2^ = 86.04) (Supplementary Figure 5).

On subgroup analysis based on the time horizon of the study (n = 23)^20, 26-28, 44-46, 48-50, 52-58, 60, 61^, EPS therapy is significantly cost-effective compared with other lipid-lowering therapeutic agents or placebo with an INBp of $4,571 (746 to 8,395) but with substantial heterogeneity (I^2^ = 85.50). However, among studies with a non-lifetime horizon, EPS therapy is not significantly cost-effective with an INBp of $381 (−10,332 to 11,093)^51, 59^ and no heterogeneity (I^2^ = 0.0) (Supplementary Figure 6).

Subgroup analysis based on discount rates of 3% (n = 15)^20, 28, 44, 49, 52, 53, 55-58, 60^ with INBp $1,879 (−4,547 to 8,305), 3.5% (n = 5) ^46-48, 51, 58^ with INBp $735 (−1,309 to 2,778) and 5% (n = 2)^26, 27^ with INBp $2,342 (−9,770 to 14,453) showed EPS therapy is not significantly cost effective compared to other lipid-lowering therapeutic agents or placebo. The 3% discount rate subgroup had considerable heterogeneity (I^2^ = 86.27), but the 3.5% and 5% discount rates subgroup had no heterogeneity (I^2^ = 0.0). However, with a discount rate of 4% (n = 3)^50, 54, 61^, EPS therapy is significantly cost-effective with an INBp of $12,254 (4,448 to 20,060) and considerable heterogeneity (I^2^ = 93.52) (Supplementary Figure 7).

EPS therapy is significantly cost-effective for primary prevention (n = 7)^27, 46, 47, 50, 51, 58^ with an INBp of $4,814 (2,523 to 7,106) and no heterogeneity (I^2^ = 0.0). However, the pooled INBs showed that EPS therapy is not significantly cost-effective for secondary prevention (n = 15)^20, 28, 44, 48, 49, 53-57, 59, 60^ with an INBp of $2,088 (−3,282 to 7,457) and considerable heterogeneity (I^2^ = 87.32). Similarly, for primary and secondary prevention together (n = 3)^26, 52, 61^ INBp $15,257 (−10,035 to 40,550) with considerable heterogeneity (I^2^ = 94.32) showed EPS therapy is not significantly cost-effective (Supplementary Figure 8).

The thresholds used in the comparisons ranged from PPP adjusted $13,218 to $2,05,198. Based on the median threshold of $50,000, the mean INBp among both the subgroups, viz., thresholds of >50,000 $ (n = 13)^20, 27, 44, 46, 53, 54, 56-58, 60, 61^ with INBp $7,398 (−1,657 to 16,452) and considerable heterogeneity (I^2^ = 89.62) and thresholds of <50,000 $ (n = 12) ^26, 28, 45, 48-52, 55, 58, 59^, INBp $1,886 (−2 to 3,775) with moderate heterogeneity (I^2^ = 36.66) showed EPS therapy is not cost effective compared with other lipid-lowering therapeutic agents or placebo, but without statistical significance (Supplementary Figure 9).

On subgroup analysis based on scenario, the mean INBp showed EPS therapy is not significantly cost effective compared with other lipid-lowering therapeutic agents or placebo for scenarios two (n = 6)^44, 53, 57, 60^ with an INBp of $-522 (−59,813 to 58,769) and considerable heterogeneity (I^2^ = 93.99). Under scenarios three (n = 3)^58, 61^ with INBp $14,398 (−12,225 to 41,020) and with considerable heterogeneity (I^2^ = 87.92) and four (n = 5)^28, 48, 50, 56, 59^ with INBp $2,876 (−568 to 6,320) with moderate heterogeneity (I^2^ = 48.81), EPS therapy was not significantly cost effective compared with other lipid-lowering therapeutic agents or placebo. However, under scenario five (n = 11)^20, 26-28, 45, 46, 49, 51, 52, 54, 55^, EPS therapy was significantly cost effective with INBs of $3,102 (599 to 5,606) and substantial heterogeneity (I^2^ = 56.67) (Supplementary Figure 10).

Further subgroup analysis was conducted to explore the heterogeneity observed when EPS therapy was compared with statin monotherapy. We found that compared with statin monotherapy, EPS therapy is significantly cost-effective for primary prevention (n = 5)^27, 46, 50, 58^ with an INBp $4,992 (2,659 to 7,326) and no heterogeneity (I^2^ = 0.0). Also, from a payers’ perspective (n = 3) ^27, 48, 52^ with INBp $2,029 (72 to 3,987) and less heterogeneity (I^2^ = 28.65) EPS therapy is significantly cost-effective compared with statin monotherapy. However, EPS therapy was cost-effective but without statistical significance among the subgroups of HICs (n = 15) with INBp $2,574 (−1,341 to 6,488) and with substantial heterogeneity (I^2^ = 86.83), UMICs (n = 3) with INBp $1,140 (−18,959 to 21,239) with no heterogeneity (I^2^ = 0.0). Similar results were also observed in healthcare perspective (n = 12)^26, 28, 44, 47, 53-55, 58-61^ with INBp $-323 (−12,568 to 11,922) and substantial heterogeneity (I^2^ = 86.40), societal perspective (n = 3) ^28, 49, 50^ with INBp $1,961 (−4,300 to 8,222) with substantial heterogeneity (I^2^ = 86.04), primary and secondary prevention together (n = 3) ^26, 52, 61^ with INBp $15,257 (−10,035 to 40,550) with substantial heterogeneity (I^2^ = 94.32) and lifetime horizon (n = 17)^26-28, 44, 46, 48, 49, 52-55, 58, 60, 61^ with INBp $2,589 (−1,293 to 6,471) with substantial heterogeneity (I^2^ = 84.95). For secondary prevention (n = 10)^28, 44, 48, 49, 53-55, 59, 60^ INBp $-2,303 (−7,728 to 3,123) showed that EPS therapy is not cost-effective compared to statin monotherapy but without statistical significance and substantial heterogeneity (I^2^ = 84.65).

### Certainty of evidence

The GRADE quality assessment revealed low confidence in the overall evidence of the cost-effectiveness of EPS therapy when compared with other lipid-lowering therapeutic agents or placebo. We found moderate confidence in results for HICs and a low level of confidence in primary prevention. However, the confidence of results from a payer perspective and lifetime horizon is very low. Considering EPS therapy compared with statin monotherapy, we have moderate confidence in the results observed for primary prevention and from the payers’ perspective, as detailed in Appendix IV.

## Discussion

The current study synthesized the cost-effectiveness of Ezetimibe with statin compared to other lipid-lowering therapeutic agents or placebo using systematic review and meta-analysis of cost-utility studies. Economic evaluation studies are difficult to synthesise due to the differences in economic parameters, income thresholds, study perspectives, costs. Many studies that report cost-effectiveness ignore the CI of the ICER point estimates. To address these issues, we tried to standardise data extraction and preprocessing from various published studies to produce a pooled INB with CI.

The pooled INBs from 25 comparisons identified from 21 studies for the meta-analysis show that EPS therapy is more cost-effective than other lipid-lowering therapeutic agents such as statins, PCSK9i, placebo, or no treatment. Subgroup analysis strengthened the robustness of our findings. We conducted a subgroup analysis to understand the considerable heterogeneity. EPS therapy is significantly cost-effective compared to PCSK9i plus statin and PCSK9i plus Ezetimibe with statins. A plethora of evidence has shown that the main reasons for the underuse of statins in LMICs and UMCs by eligible patients with established CVD^63, 64^ were lack of availability, accessibility, and affordability. The subgroup analysis also revealed that EPS therapy is cost-effective against therapeutic agents or placebo in HICs, with payers’ perspective, lifetime horizon, and primary prevention. However, the results lose their robustness and become not significantly cost-effective for HICs and lifetime horizons when we limit the comparator to statin monotherapy alone in further subgroup analysis.

Previous systematic review by Marquina et al. ^65^, had indicated that Ezetimibe was cost-effective in 62.5% of the included studies, and Suh et al. ^66^, indicated that Ezetimibe was cost-effective for stain intolerant patients with chronic kidney disease. With the current evidence, we conclude EPS therapy is not cost-effective for secondary prevention, similar to previously published studies that suggested that PCSK9i may become cost-effective for secondary prevention ^61^ only if the WTP threshold is increased or if the drugs cost is lowered ^44, 60^.

For Ezetimibe, the main cost-effectiveness drivers were baseline cardiovascular risks ^26, 45, 46, 48, 51, 55^, cost of the drug ^44, 52, 54, 60^, treatment effects related to cardiovascular and non-cardiovascular mortality ^28, 52^, and time horizon ^44, 55, 59^. For some other studies, the cost-effectiveness of EPS therapy compared with statin monotherapy was subject to certain conditions. Davies et al. ^52^ reported that EPS therapy was cost-effective for secondary prevention of CHD and stroke. Also, the study reported that for primary prevention of CHD and stroke in patients whose LDL-C levels were > 100 mg/dL and in patients with diabetes, Ezetimibe becomes cost-effective only if we consideri a 90% cost reduction.. Soini et al. ^49^ showed that EPS therapy is cost-effective only in specific sub-populations of men and diabetic women. A study by Ara et al. ^46^ reported that EPS therapy becomes cost-effective in the UK health system when using a threshold of £30,000 per QALY instead of the £20,000 per QALY value. Similarly, Almalki et al. ^55^ reported that EPS therapy was not cost-effective for 5 and 10 year models but became a cost-effective treatment when a lifetime horizon is used.

The drug price and WTP per additional QALY play an important role in determining the cost-effectiveness of EPS therapy. The differences observed in reported outcomes and the high heterogeneity estimated among the studies may be the result of changes in drug prices or cost estimations under different study perspectives. This necessitates the need for context-specific future research in primary economic evaluations for EPS therapy compared with different lipid-lowering therapeutic agents with a more accurate estimate of costs. More studies from LMICs and societal perspectives are also needed for the generalisability of results.

The limitation of our study is that the comprehensiveness of this cost-effectiveness results is arguable since most clinical data comes from western countries, which are set in HICs, which determine the cost of long-term treatment and prescriptions. Only a small percentage comes from Asian studies and none from Africa, Australia, or South America, indicating the need for more studies using LMICs and societal perspectives. A Majority of the studies have not included indirect costs nor. And none captured the real-world burden of CVD or considered out-of-pocket expenditure and caregiver time and costs. The use of surrogate markers even when clinical endpoints areis recommended^67^, t shows lack of clinical trials describing complex outcomes and have increased the uncertainties in the models. The use of clinical trials to model event rates could lead to an underestimation of the baseline risk and the treatment effect, as shown in Lindh et al. ^68^. The mean age reported in majority of the studes was above 60 years.. Although LDL-C poses a cumulative risk, lowering LDL-C levels does not always result in a reduction in cardiovascular risk, and prolonged exposure to lower LDL-C from an early point in life substantially reduces the risk of CHD ^69^. Given the focus on populations with established CVD, most studies did not examine the effects of treatment initiation at different ages. Moreover, some models used statin trial data to model baseline cardiovascular risk, while others used local demographic data.

Another limitation is that we used funnel plots to examine publication bias because we had no specific measures for non-normally distributed INB. We used the GRADE approach to assess the outcome quality because there were no specific GRADE guidelines for cost-utility studies, and the current approach had its limitations ^70^. We could not discern a clear trend regarding whether using data from clinical trials instead of observational studies in terms of baseline risk and treatment effect improves the cost-effectiveness results.. The structural variation among studies could also raise the overall conclusion’s uncertainty. Many of thesemodels are not publicly available which limits the ability to compare between settings and countries ^71^. Only four studies ^44, 53, 59, 60^ included adverse events. Adherence was not modelled in any studies, even when lower adherence rates have been shown to result in poorer health outcomes and higher costs for healthcare systems time after time^72^When extrapolated to other healthcare contexts, the generalisability of these results should be done with careful consideration. Further, due to limited informationprovided, the,costs for co-morbidities and gender differences could not be explored.

## CONCLUSION

This systematic review and meta-analysis concluded that EPS therapy is a significant cost-effective option compared to other lipid-lowering therapeutic agents or placebo. The subgroup analysis supported the findings in HICs, from payers’ perspective, for primary prevention and for the lifetime horizon. However, the robustness of the results is lost for HICs and lifetime horizon when EPS therapy is compared with statin monotherapy alone, where it is not significantly cost-effective. The GRADE quality assessment revealed moderate confidence in the results observed in HICs for EPS therapy compared to other lipid-lowering therapeutic agents or placebo and statin monotherapy. The majority of the studies were from HICs and undertook a healthcare perspective, highlighting a lacuna in evidence to be filled from a societal perspective and LMICs.

## Data Availability

All relevant data are within the manuscript and its Supporting Information files.

## Acknowledgements

None

## References

1. Karr S. Epidemiology and management of hyperlipidemia. The American Journal of Managed Care 2017;23(9 Suppl):S139–S148.

2. Navarese EP, Robinson JG, Kowalewski M, Kolodziejczak M, Andreotti F, Bliden K, et al. Association Between Baseline LDL-C Level and Total and Cardiovascular Mortality After LDL-C Lowering: A Systematic Review and Meta-analysis. JAMA 2018;319(15):1566–1579.

3. Stone NJ, Robinson JG, Lichtenstein AH, Bairey Merz CN, Blum CB, Eckel RH, et al. 2013 ACC/AHA guideline on the treatment of blood cholesterol to reduce atherosclerotic cardiovascular risk in adults: a report of the American College of Cardiology/American Heart Association Task Force on Practice Guidelines. Journal of the American College of Cardiology 2014;63(25 Part B):2889–2934.

4. Organisation Wh. Cardiovascular diseases (CVDs). In: WHO.

5. Efficacy and safety of LDL-lowering therapy among men and women: meta-analysis of individual data from 174 000 participants in 27 randomised trials. The Lancet 2015;385(9976):1397–1405.

6. Catapano AL, Graham I, De Backer G, Wiklund O, Chapman MJ, Drexel H, et al. 2016 ESC/EAS guidelines for the management of dyslipidaemias: the task force for the management of dyslipidaemias of the European Society of Cardiology (ESC) and European Atherosclerosis Society (EAS) developed with the special contribution of the European Association for Cardiovascular Prevention & Rehabilitation (EACPR). Atherosclerosis 2016;253:281–344.

7. Grundy SM, Stone NJ, Bailey AL, Beam C, Birtcher KK, Blumenthal RS, et al. 2018 AHA/ACC/AACVPR/AAPA/ABC/ACPM/ADA/AGS/APhA/ASPC/NLA/PCNA guideline on the management of blood cholesterol: executive summary: a report of the American College of Cardiology/American Heart Association Task Force on Clinical Practice Guidelines. Journal of the American College of Cardiology 2019;73(24):3168–3209.

8. Mihaylova B, Emberson J, Blackwell L, Keech A, Simes J, Barnes E, et al. The effects of lowering LDL cholesterol with statin therapy in people at low risk of vascular disease: meta-analysis of individual data from 27 randomised trials. Lancet (London, England) 2012;380(9841):581–590.

9. Bavry AA, Mood GR, Kumbhani DJ, Borek PP, Askari AT, Bhatt DL. Long-Term Benefit of Statin Therapy Initiated??during Hospitalization for??an??Acute??Coronary Syndrome: A Systematic Review of Randomized Trials. American Journal of Cardiovascular Drugs 2007;7(2):135–141.

10. Brugts JJ, Yetgin T, Hoeks SE, Gotto AM, Shepherd J, Westendorp RGJ, et al. The benefits of statins in people without established cardiovascular disease but with cardiovascular risk factors: meta-analysis of randomised controlled trials. BMJ 2009;338(jun30 1):b2376–b2376.

11. Boekholdt SM, Hovingh GK, Mora S, Arsenault BJ, Amarenco P, Pedersen TR, et al. Very low levels of atherogenic lipoproteins and the risk for cardiovascular events: a meta-analysis of statin trials. Journal of the American College of Cardiology 2014;64(5):485–494.

12. Tataronis GR. Statin-Related Adverse Events: A Meta-Analysis. Clinical Therapeutics 2006;28(1).

13. Kosoglou T, Meyer I, Veltri EP, Statkevich P, Yang B, Zhu Y, et al. Pharmacodynamic interaction between the new selective cholesterol absorption inhibitor ezetimibe and simvastatin. British journal of clinical pharmacology 2002;54(3):309–319.

14. Sudhop T, Lütjohann D, Kodal A, Igel M, Tribble DL, Shah S, et al. Inhibition of intestinal cholesterol absorption by ezetimibe in humans. Circulation 2002;106(15):1943–1948.

15. Cannon CP, Blazing MA, Giugliano RP, McCagg A, White JA, Theroux P, et al. Ezetimibe added to statin therapy after acute coronary syndromes. New England Journal of Medicine 2015;372(25):2387–2397.

16. Morrone D, Weintraub WS, Toth PP, Hanson ME, Lowe RS, Lin J, et al. Lipid-altering efficacy of ezetimibe plus statin and statin monotherapy and identification of factors associated with treatment response: a pooled analysis of over 21,000 subjects from 27 clinical trials. Atherosclerosis 2012;223(2):251–261.

17. Claxton K, Sculpher M, McCabe C, Briggs A, Akehurst R, Buxton M, et al. Probabilistic sensitivity analysis for NICE technology assessment: not an optional extra. Health economics 2005;14(4):339–347.

18. Lloyd-Jones DM, Morris PB, Ballantyne CM, Birtcher KK, Daly DD, DePalma SM, et al. 2017 focused update of the 2016 ACC expert consensus decision pathway on the role of non-statin therapies for LDL-cholesterol lowering in the management of atherosclerotic cardiovascular disease risk: a report of the American College of Cardiology Task Force on Expert Consensus Decision Pathways. Journal of the American College of Cardiology 2017;70(14):1785–1822.

19. Orringer CE, Jacobson TA, Saseen JJ, Brown AS, Gotto AM, Ross JL, et al. Update on the use of PCSK9 inhibitors in adults: Recommendations from an Expert Panel of the National Lipid Association. Journal of clinical lipidology 2017;11(4):880–890.

20. Ulf Landmesser PL, Emil Hagstróm, Ben van Hout,, Guillermo Villa PP-R, Jorge Arellano, Maria Eriksson Svensson, Mahendra Sibartie, and Gregg C Fonarow. Cost-effectiveness of PCSK9 inhibition with evolocumab in patients with a history of myocardial infarction in Sweden. European heart journal Quality of care & clinical outcomes 2020.

21. Robinson JG, Jayanna MB, Brown AS, Aspry K, Orringer C, Gill EA, et al. Enhancing the value of PCSK9 monoclonal antibodies by identifying patients most likely to benefit. A consensus statement from the National Lipid Association. Journal of clinical lipidology 2019;13(4):525–537.

22. Chaiyasothi T, Nathisuwan S, Dilokthornsakul P, Vathesatogkit P, Thakkinstian A, Reid C, et al. Effects of non-statin lipid-modifying agents on cardiovascular morbidity and mortality among statin-treated patients: a systematic review and network meta-analysis. Frontiers in pharmacology 2019;10:547.

23. Bagepally BS, Sasidharan A. Incremental net benefit of lipid-lowering therapy with PCSK9 inhibitors: a systematic review and meta-analysis of cost-utility studies. European Journal of Clinical Pharmacology 2021.

24. Arbel R, Hammerman A, Azuri J. Usefulness of Ezetimibe Versus Evolocumab as Add-On Therapy for Secondary Prevention of Cardiovascular Events in Patients With Type 2 Diabetes Mellitus. Am J Cardiol 2019;123(8):1273–1276.

25. Qamar A, Giugliano RP, Bohula EA, Park J-G, Jarolim P, Murphy SA, et al. Biomarkers and Clinical Cardiovascular Outcomes With Ezetimibe in the IMPROVE-IT Trial. Journal of the American College of Cardiology 2019;74(8):1057–1068.

26. Kohli M, Attard C, Lam A, Huse D, Cook J, Bourgault C, et al. Cost effectiveness of adding ezetimibe to atorvastatin therapy in patients not at cholesterol treatment goal in Canada. Pharmacoeconomics 2006;24(8):815–830.

27. Laires PA, Ejzykowicz F, Hsu T-Y, Ambegaonkar B, Davies G. Cost-effectiveness of adding ezetimibe to atorvastatin vs switching to rosuvastatin therapy in Portugal. J Med Econ 2015;18(8):565–572.

28. Kongpakwattana K, Ademi Z, Chaiyasothi T, Nathisuwan S, Zomer E, Liew D, et al. Cost-Effectiveness Analysis of Non-Statin Lipid-Modifying Agents for Secondary Cardiovascular Disease Prevention Among Statin-Treated Patients in Thailand. PharmacoEconomics 2019;37(10):1277–1286.

29. Moher D SL, PRISMA-P Group,. Preferred reporting items for systematic review and meta-analysis protocols (PRISMA-P) 2015 statement.. Syst Rev 2015;4(1):1.

30. Registry C. Center for the Evaluation of Value and Risk in Health. In.

31. Ouzzani M HH, Fedorowicz Z, Elmagarmid A. Rayyan-a web and mobile app for systematic reviews. Syst Rev 2016;5(1):210.

32. Rohatgi A. WebPlotDigitizer. In. USA; 2021.

33. Bagepally BS, Gurav YK, Anothaisintawee T, Youngkong S, Chaikledkaew U, Thakkinstian A. Cost Utility of Sodium-Glucose Cotransporter 2 Inhibitors in the Treatment of Metformin Monotherapy Failed Type 2 Diabetes Patients: A Systematic Review and Meta-Analysis. Value Health 2019;22(12):1458–1469.

34. Paulden M. Why it’s Time to Abandon the ICER. Pharmacoeconomics 2020;38(8):781–784.

35. O’Mahony JF. The Limitations of Icers In Screening Interventions and The Relative Net Benefit Alternative. Value in Health 2015;18(7).

36. Bagepally BS, Chaikledkaew U, Gurav YK, Anothaisintawee T, Youngkong S, Chaiyakunapruk N, et al. Glucagon-like peptide 1 agonists for treatment of patients with type 2 diabetes who fail metformin monotherapy: systematic review and meta-analysis of economic evaluation studies. BMJ Open Diabetes Res Care 2020;8(1).

37. Crespo C, Monleon A, Díaz W, Ríos M. Comparative efficiency research (COMER): meta-analysis of cost-effectiveness studies. BMC medical research methodology 2014;14(1):1–9.

38. Bank TW. World Bank Country and Lending Groups – World Bank Data Help Desk. In; 2021.

39. >Corporation. M. Microsoft Excel [Internet]. In; 2018.

40. StataCorp. Stata Statistical Software: Release 16. In. 16 ed: College Station, TX: StataCorp LLC.; 2019.

41. Adarkwah CCvGP, Hiligsmann M, Evers SMAA. Risk of bias in model-based economic evaluations: the ECOBIAS checklist. Expert Review of Pharmacoeconomics & Outcomes Research 2016;16(4):513–523.

42. Guyatt G, Oxman AD, Akl EA, Kunz R, Vist G, Brozek J, et al. GRADE guidelines: 1. Introduction—GRADE evidence profiles and summary of findings tables. Journal of Clinical Epidemiology 2011;64(4):383–394.

43. Hultcrantz M, Rind D, Akl EA, Treweek S, Mustafa RA, Iorio A, et al. The GRADE Working Group clarifies the construct of certainty of evidence. Journal of Clinical Epidemiology 2017;87:4–13.

44. Kazi DS, Moran AE, Coxson PG, Penko J, Ollendorf DA, Pearson SD, et al. Cost-effectiveness of PCSK9 Inhibitor Therapy in Patients With Heterozygous Familial Hypercholesterolemia or Atherosclerotic Cardiovascular Disease. JAMA 2016;316(7):743.

45. Ara R, Pandor A, Tumur I, Paisley S, Duenas A, Williams R, et al. Cost effectiveness of ezetimibe in patients with cardiovascular disease and statin intolerance or contraindications: a Markov model. Am J Cardiovasc Drugs 2008;8(6):419–427.

46. Ara R, Pandor A, Tumur I, Paisley S, Duenas A, Williams R, et al. Estimating the health benefits and costs associated with ezetimibe coadministered with statin therapy compared with higher dose statin monotherapy in patients with established cardiovascular disease: Results of a Markov model for UK costs using data registries. Clin Ther 2008;30(8):1508–1523.

47. Ara R, Tumur I, Pandor A, Duenas A, Williams R, Wilkinson A, et al. Ezetimibe for the treatment of hypercholesterolaemia: A systematic review and economic evaluation. Health Technol Assess 2008;12(21):1–92.

48. Reckless J, Davies G, Tunceli K, Hu XH, Brudi P. Projected cost-effectiveness of ezetimibe/simvastatin compared with doubling the statin dose in the United Kingdom: Findings from the INFORCE study. Value Health 2010;13(6):726–734.

49. Soini EJO, Davies G, Martikainen JA, Hu HX, Tunceli K, Niskanen L. Population-based health-economic evaluation of the secondary prevention of coronary heart disease in Finland. Curr Med Res Opin 2010;26(1):25–36.

50. F. van Nooten Gmd, J. W. Jukema, A. H. Liem, E. Yap, X. H. Hu. Economic evaluation of ezetimibe combined with simvastatin for the treatment of primary hypercholesterolaemia. Netherlands heart journal : monthly journal of the Netherlands Society of Cardiology and the Netherlands Heart Foundation 2011;19(2):61–7.

51. Mihaylova B S, Herrington W, Lozano-Kühne J, Kent S, Emberson J, Reith C, Haynes R, Cass A, Craig J, Gray A, Collins R, Landray M C B., Cost-effectiveness of Simvastatin plus Ezetimibe for Cardiovascular Prevention in CKD: Results of the Study of Heart and Renal Protection (SHARP). American journal of kidney diseases : the official journal of the National Kidney Foundation 2016;67(4):576–84.

52. Davies GM, Vyas A, Baxter CA. Economic evaluation of ezetimibe treatment in combination with statin therapy in the United States. J Med Econ 2017;20(7):723–731.

53. Kazi DS, Penko J, Coxson PG, Moran AE, Ollendorf DA, Tice JA, et al. Updated cost-effectiveness analysis of PCSK9 inhibitors based on the results of the FOURIER trial. JAMA 2017;318(8):748–750.

54. Stam-Slob MC, van der Graaf Y, Greving JP, Dorresteijn JAN, Visseren FLJ. Cost-effectiveness of intensifying lipid-lowering therapy with statins based on individual absolute benefit in coronary artery disease patients. Journal of the American Heart Association 2017;6(2).

55. Almalki ZS, Guo JJ, Alahmari A, Alotaibi N, Thaibah H. Cost-Effectiveness of Simvastatin Plus Ezetimibe for Cardiovascular Prevention in Patients With a History of Acute Coronary Syndrome: Analysis of Results of the IMPROVE-IT Trial. Heart, Lung and Circulation 2018;27(6):656–665.

56. Stam-Slob MC, van der Graaf Y, de Boer A, Greving JP, Visseren FLJ. Cost-effectiveness of PCSK9 inhibition in addition to standard lipid-lowering therapy in patients at high risk for vascular disease. Int J Cardiol 2018;253:148–154.

57. Dressel A, Schmidt B, Schmidt N, Laufs U, Fath F, Chapman MJ, et al. Cost effectiveness of lifelong therapy with PCSK9 inhibitors for lowering cardiovascular events in patients with stable coronary artery disease: Insights from the Ludwigshafen Risk and Cardiovascular Health cohort. Vasc Pharmacol 2019;120.

58. Iryna Schlackow SK, William Herrington3, Jonathan Emberson, Richard Haynes, Christina Reith, Rory Collins, Martin J. Landray, Alastair Gray, Colin Baigent and Borislava Mihaylova Cost-effectiveness of lipid lowering with statins and ezetimibe in chronic kidney disease. Kidney International 2019;96(1):170–179.

59. Yang H, Li N, Zhou Y, Xiao Z, Tian H, Hu M, et al. Cost-effectiveness analysis of Ezetimibe as the add-on treatment to moderate-dose rosuvastatin versus high-dose rosuvastatin in the secondary prevention of cardiovascular diseases in China: A Markov model analysis. Drug Des Dev Ther 2020;14:157–165.

60. Kazi DS, Penko J, Coxson PG, Guzman D, Wei PC, Bibbins-Domingo K. Cost-Effectiveness of Alirocumab: A Just-in-Time Analysis Based on the ODYSSEY Outcomes Trial. Annals of Internal Medicine 2019;170(4):221.

61. Korman M, Wisløff T. Modelling the cost-effectiveness of PCSK9 inhibitors vs. ezetimibe through LDL-C reductions in a Norwegian setting. European Heart Journal - Cardiovascular Pharmacotherapy 2018;4(1):15–22.

62. Adarkwah CC, van Gils PF, Hiligsmann M, Evers SM. Risk of bias in model-based economic evaluations: the ECOBIAS checklist. Expert Rev Pharmacoecon Outcomes Res 2016;16(4):513–23.

63. Husain MJ, Datta BK, Kostova D, Joseph KT, Asma S, Richter P, et al. Access to cardiovascular disease and hypertension medicines in developing countries: an analysis of essential medicine Lists, price, availability, and affordability. Journal of the American Heart Association 2020;9(9):e015302.

64. Wirtz VJ, Kaplan WA, Kwan GF, Laing RO. Access to medications for cardiovascular diseases in low-and middle-income countries. Circulation 2016;133(21):2076–2085.

65. Marquina C, Zomer E, Vargas-Torres S, Zoungas S, Ofori-Asenso R, Liew D, et al. Novel Treatment Strategies for Secondary Prevention of Cardiovascular Disease: A Systematic Review of Cost-Effectiveness. PharmacoEconomics 2020;38(10):1095–1113.

66. Suh D-C, Griggs SK, Henderson ER, Lee S-M, Park T. Comparative effectiveness of lipid-lowering treatments to reduce cardiovascular disease. Expert Review of Pharmacoeconomics & Outcomes Research 2018;18(1):51–69.

67. (NICE) NIfHaCE. Guide to the methods of technology appraisal 2013. In; 04 April 2013. p. 94.

68. Lindh M, Banefelt J, Fox KM, Hallberg S, Tai M-H, Eriksson M, et al. Cardiovascular event rates in a high atherosclerotic cardiovascular disease risk population: estimates from Swedish population-based register data. European Heart Journal -Quality of Care and Clinical Outcomes 2019;5(3):225–232.

69. Ference BA, Yoo W, Alesh I, Mahajan N, Mirowska KK, Mewada A, et al. Effect of long-term exposure to lower low-density lipoprotein cholesterol beginning early in life on the risk of coronary heart disease: a Mendelian randomization analysis. Journal of the American College of Cardiology 2012;60(25):2631–2639.

70. Brozek JL, Canelo-Aybar C, Akl EA, Bowen JM, Bucher J, Chiu WA, et al. GRADE Guidelines 30: the GRADE approach to assessing the certainty of modeled evidence— An overview in the context of health decision-making. Journal of Clinical Epidemiology 2021;129:138–150.

71. Wu EQ, Zhou Z-Y, Xie J, Metallo C, Thokala P. Transparency in Health Economic Modeling: Options, Issues and Potential Solutions. PharmacoEconomics 2019;37(11):1349–1354.

72. Cutler RL, Fernandez-Llimos F, Frommer M, Benrimoj C, Garcia-Cardenas V. Economic impact of medication non-adherence by disease groups: a systematic review. BMJ Open 2018;8(1):e016982.

